# g-CXR-Net: A Graphic Application for the Rapid Recognition of SARS-CoV-2 from Chest X-Rays

**DOI:** 10.1101/2021.06.06.21258428

**Authors:** Domenico L. Gatti

## Abstract

g-CXR-Net is a graphic application for the rapid recognition of SARS-CoV-2 from Antero/Posterior chest X-rays. It employs the Artificial Intelligence engine of CXR-Net (<u>arXiv:2103.00087</u>) to generate masks of the lungs overlapping the heart and large vasa, probabilities for Covid *vs*. non-Covid assignment, and high resolution heat maps that identify the SARS associated lung regions.

## 1. Introduction

A critical step in the fight against COVID-19 is the rapid recognition of patients affected by Severe Acute Respiratory Syndrome CoronaVirus 2 (SARS-CoV-2). The majority of COVID-19 patients with pneumonia present bilateral abnormalities mostly in the form of ground-glass opacities and consolidations in computed tomography (CT) and/or chest X-ray (CXR) images [1]. While CTs provide greater diagnostic accuracy, CXRs are more readily available and enable rapid triaging of patients. However, correct interpretation of CXRs is often challenging, and new methods of Machine Learning (ML) and Artificial Intelligence (AI) aim to provide assistance in this task [2]. As part of this effort my group has developed CXR-Net [3], a Convolutional Neural Network (CNN) two-module pipeline for SARS-CoV-2 detection. Module I was trained on datasets of Antero/Posterior CXRs with radiologist annotated lung contours to generate accurate masks of the lungs that overlap the heart and large vasa, and are minimally influenced by regions of consolidation or other texture alterations from underlying pathologies. Module II is a hybrid convnet in which the first convolutional layer with learned coefficients is replaced by a layer with fixed coefficients provided by the Wavelet Scattering Transform (WST). A particular advantage of this hybrid net is the removal of network instabilities associated with adversarial images (noise or small deformations in the input images that are visually insignificant, but that the network does not reduce correctly, leading to incorrect classification). This net converges more rapidly than an end-to-end learned architecture, does not suffer from vanishing or exploding gradients, and prevents overfitting, leading to better generalization. Module II takes as inputs the patients’ CXRs and corresponding lung masks calculated by Module I, and produces as outputs a class assignment (Covid *vs*. non-Covid) and high resolution heat maps that identify the SARS associated lung regions. CXR-Net was implemented using *Keras* [4] deep learning library running on top of *TensorFlow* 2.2 [5], and is publicly available at https://github.com/dgattiwsu/CXR-Net.

Here we introduce g-CXR-Net, a graphic application based on the CXR-Net segmentation and classification engine. g-CXR-Net is publicly available at https://github.com/dgattiwsu/g-CXR-Net. It adopts the Python *<u>tkinter</u>* package as the standard interface to the Tk GUI toolkit (https://www.tcl.tk/), and thus is compatible with most Unix platforms, as well as Windows systems.

## 2. g-CXR-Net architecture

We recall here briefly the key features of CXR-Net, the AI engine at the heart of g-CXR-Net.

CXR-Net Module I is a new type of segmentation engine that offers some advantages with respect to the traditional encoder-decoder architecture, as its layers contain no pooling or up-sampling operations, and therefore the spatial dimensions of the feature maps at each layer remain unchanged with respect to those of the input images and of the segmentation masks used as labels or predicted by the network. For this reason, Module I can process images of any size and shape without changing layers size and operations.

CXR-Net Module II is a novel type of convolutional classification network for biomedical images, in which the initial convolutional layers with learned coefficients are replaced by the Wavelet Scattering Transform (WST) of the input image, with fixed coefficients determined only by the scales and rotations of the analytical wavelet used in the transform [6]. The use of the WST as an image pre-processing step is equivalent to a process of transfer learning in which the input image is first passed through a CNN pretrained on a very large number of unrelated images, in order to provide an initial feature map that generalizes well also to the specific case at hand. The WST block of CXR-Net Module II produces a feature map that is passed on to a CNN with the same general architecture as Module I, so that all intermediate feature maps maintain the same dimensions as the initial input image. This type of hybrid network achieves excellent classification performance with only a small number of learned coefficients.

In fact, CXR-Net Module II requires the refinement of only ∼21,000 parameters for images of dimensions 300 × 340. For this reason, the models derived from 6 independent cross-validated training runs of Module II were combined without averaging into a single *ensemble model* that uses only ∼123,000 parameters.

## 3. g-CXR-Net GUI

Upon download from its *github* repository g-CXR-Net expands into a tree of directories and files as shown in Fig. 1. The GUI, driven by the python script *g_CXR_Net*.*py*, can be launched from the command line, invoking the python executable in the *bin* directory of the virtual environment where all required libraries (i.e., *Tensorflow, Keras, Kymatio*) are installed, or from a linked icon. When g-CXR-Net is used for the first time to process a CXR image, the underlying CXR-Net engine is built *ex novo*, and its refined coefficients are loaded from the files in the *models* folder (Fig. 1). However, once both the segmentation and classification models of CXR-Net are in memory, any additional CXR image is processed in just a few seconds on a standard laptop or desktop computer, without the need for GPUs. The GUI displays a single panel (Fig. 2) with buttons assigned to different tasks. These are: a) select a directory tree for sources and target files different from the default one created by the application, b) select CXR images in DCM format to be processed, c) update the selections of folders and image files, and display results (Fig. 2, bottom left window), d) calculate the lung masks, e) calculate the scores of each CXR as the probability of being *Covid* or *non-Covid*, f) calculate heat maps justifying the class assignment and the classification scores, g) display CXRs and the corresponding calculated lung masks (Fig. 2, top right window), h) display the heat maps (Fig. 2, bottom right window), i) quit.

**Fig. 1.**
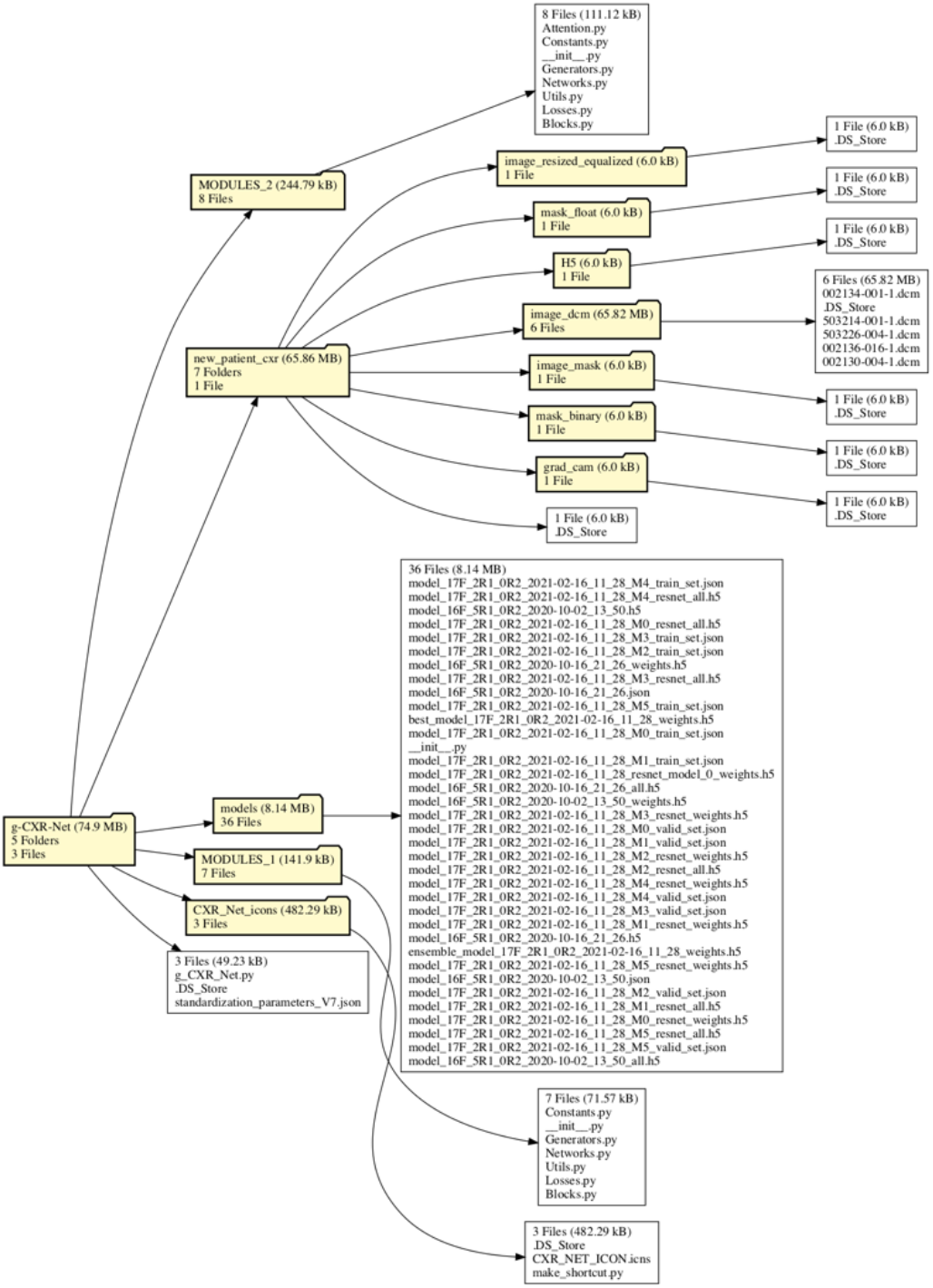
g-CXR-Net tree. *MODULES_I* and *MODULES_II* folders contain all the modules necessary to build the underlying AI engine of CXR-Net. Trained CXR-Net models are in the *models* folder. The *new_patient_cxr* folder contains both the source directory for CXR images in DCM format, and the target directories where *g-CXR-Net* stores CXR images converted from DCM to PNG format, intermediate files in H5 format used for classification, the binary and floating point lung masks generated by CXR-Net Module I, and the heat maps, displaying the lung regions with potential SARS-CoV-2 lesions, generated by CXR-Net Module II. The folder CXR_Net_icons contains an example icon for the application, and a python script that can be used to generate an icon link to *g_CXR_Net*.*py* anywhere in the OS tree.

**Fig. 2.**
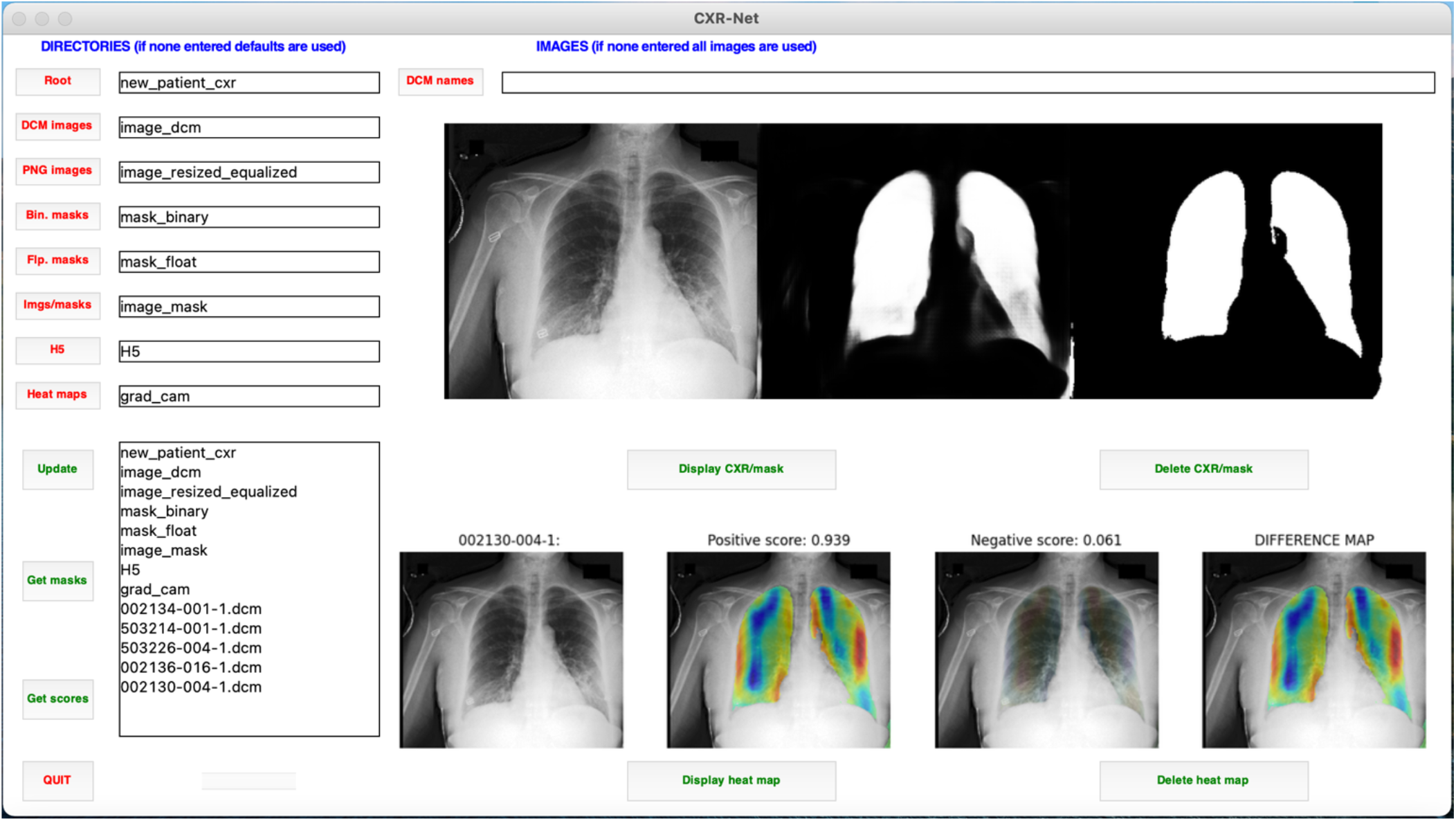
g-CXR-Net GUI. Buttons are associated with different actions: a) select a directory tree for sources and targets different from the default one, b) select CXR images to be processed, c) update selections, d) calculate lung masks, e) calculate classification scores, f) calculate heat maps, g) display CXRs and corresponding floating point and binary lung masks, h) display heat maps, i) quit. Updates and task results are displayed in the text window at the lower left corner of the GUI.

g-CXR-Net uses gradient weighted class activation heat maps (Grad-CAM saliency maps, [7]) to depict visually the decisions made by CXR-Net ensemble model, so that the outcome can be evaluated critically by a radiologist. These saliency maps are generated from the two channels of the final convolutional layer of CXR-Net Module II, which are then globally averaged for direct input to a Softmax activation. Thus, the two channels contain the predictions for the CXR image probability of being Covid + or Covid –, respectively, summing up to 1. g-CXR-Net displays Grad-CAM maps from both channels, and also a difference map between the two channels (Fig. 2, bottom right window). When Module II clearly favors a Covid + or Covid – assignment, the difference map is almost identical to the map derived from the channel that produces the highest probability. When the assignment is uncertain (similar probabilities from both channels) the difference map tend to be featureless. When Module II classifies the image as Covid –, the difference maps identifies the lung regions that could still possibly be associated to a Covid form of pneumonia.

### Software

Source code for g-CXR-Net is deposited at https://github.com/dgattiwsu/g-CXR-Net. A video tutorial explaining how to install and run g-CXR-Net is available on the corresponding author’s academic web site at http://veloce.med.wayne.edu/~gatti/neural-networks/cxr-net.html.

## Data Availability

Source code for g-CXR-Net is publicly available at https://github.com/dgattiwsu/g-CXR-Net

https://github.com/dgattiwsu/CXR-Net

http://veloce.med.wayne.edu/~gatti/neural-networks/cxr-net.html

## Acknowledgements

Development of g-CXR-Net was supported by the WSU President’s Research Enhancement Program in Computational Biology.

